# Modes of contact and risk of transmission in COVID-19 among close contacts

**DOI:** 10.1101/2020.03.24.20042606

**Authors:** Lei Luo, Dan Liu, Xin-long Liao, Xian-bo Wu, Qin-long Jing, Jia-zhen Zheng, Fang-hua Liu, Shi-gui Yang, Bi Bi, Zhi-hao Li, Jian-ping Liu, Wei-qi Song, Wei Zhu, Zheng-he Wang, Xi-ru Zhang, Pei-liang Chen, Hua-min Liu, Xin Cheng, Miao-chun Cai, Qing-mei Huang, Pei Yang, Xing-fen Yang, Zhi-gang Han, Jin-ling Tang, Yu Ma, Chen Mao

## Abstract

**Background:** Rapid spread of SARS-CoV-2 in Wuhan prompted heightened surveillance in Guangzhou and elsewhere in China. Modes of contact and risk of transmission among close contacts have not been well estimated.

**Methods:** We included 4950 closes contacts from Guangzhou, and extracted data including modes of contact, laboratory testing, clinical characteristics of confirmed cases and source cases. We used logistic regression analysis to explore the risk factors associated with infection of close contacts.

**Results:** Among 4950 closes contacts, the median age was 38.0 years, and males accounted for 50.2% (2484). During quarantine period, 129 cases (2.6%) were diagnosed, with 8 asymptomatic (6.2%), 49 mild (38.0%), and 5 (3.9%) severe to critical cases. The sensitivity of throat swab was 71.32% and 92.19% at first to second PCR test. Among different modes of contact, household contacts were the most dangerous in catching with infection of COVID-19, with an incidence of 10.2%. As the increase of age for close contacts and severity of source cases, the incidence of COVID-19 presented an increasing trend from 1.8% (0-17 years) to 4.2% (60 or over years), and from 0.33% for asymptomatic, 3.3% for mild, to 6.2% for severe and critical source cases, respectively. Manifestation of expectoration in source cases was also highly associated with an increased risk of infection in their close contacts (13.6%). Secondary cases were in general clinically milder and were less likely to have common symptoms than those of source cases.

**Conclusions:** In conclusion, the proportion of asymptomatic and mild infections account for almost half of the confirmed cases among close contacts. The household contacts were the main transmission mode, and clinically more severe cases were more likely to pass the infection to their close contacts. Generally, the secondary cases were clinically milder than those of source cases.

## Introduction

In December 2019, the outbreak of Coronavirus Disease 2019 (COVID-19) caused by Severe Acute Respiratory Syndrome Coronavirus 2 (SARS-CoV-2) emerged in Wuhan, Hubei Province, China, and has now developed into a global pandemic^1^. As of 15 March, worldwide a total of 153,517 people have been infected including 5,735 deaths, with 81,048 cases and 3,204 deaths in China^2^.

The viral, epidemiological, and clinical characteristics of the disease have been documented^3-11^. However, some questions important for control of the epidemic remain outstanding^10^. For example, what is the transmissibility of the virus? What patients are more likely to spread the virus? What mode of contacts is most likely to cause transmission? What is the incidence of complete asymptomatic infection?

These questions are addressed in this follow-up study of 4,950 persons with close contact with confirmed COVID-2019 patients in Guangzhou, China.

## Methods

### Study Oversight

This is a prospective cohort study of all 4,950 persons who had a close contact (or close contacts in short) with confirmed COVID-2019 patients (or source cases in short) identified between January 13 and March 6, 2020, in Guangzhou, Guangdong Province, China. A total of 129 cases were diagnosed with 42 before quarantine and 87 during the quarantine.

### Data Sources

Close contacts include such unprotected contacts as living in the same household, face-to-face working together, sharing the same classroom, visit or stay in the same hospital ward, taking the same car or aeroplane, sharing neighbouring seats in the same train or ship as a diagnosed COVID-19 patient. It also includes giving direct care to a diagnosed patient. The full definition and whole list of forms of close contacts were showed in Appendix 1. When a COVID-19 patient was diagnosed then his or her close contacts were traced, and his or her close contacts may be locals or non-locals, if he or she had a history of travel or business. Thus, the source cases of close contacts included both local and non-local patients.

Between January 13, and March 6, 2020, 347 cases^12^ were diagnosed in Guangzhou and their 4,950 close contacts were identified and enrolled in the study. Standard questionnaires were used to collect data at the time of enrollment, which was also the start of quarantine^13^. The registration form (Appendix 2-Table a) was completed for each close contact. All close contacts were put under quarantine for 14 days from the last contact or longer for some cases if collection of samples for PCR testing was delayed. We recorded the last date of contact, the date of the start of quarantine, the date symptoms appeared, the date of each sampling, and the date of first positive PCR result. Temperature and symptoms monitoring were conducted every day and recorded in a standard form (Appendix 2-Table b).

Throat swab samples were collected and a real time RT-PCR testing performed once every two days. In one patient, the PCR testing was performed ten times as previous tests were consistently negative and has not released from quarantine. A close contact was released from quarantine if he had no symptoms and PCR testing was e negative for two consecutive samples. For those who were diagnosed with COVID-19, treatments followed and quarantine continued till recovery.

Data on demographic factors, risk factors, exposure history, mode of contact, symptoms, radiological and laboratory findings, severity of disease, treatments, and prognoses were collected on all close contacts (data form in Appendix 2). The information of source cases was also obtained through monitoring data from Guangzhou CDC. Close contacts confirmed COVID-19 (or secondary cases in short) and their source cases are individually linked (details in Appendix 2) and their relations and contact modes were determined accordingly. For the 161 source cases who did not live in Guangzhou, so we could not know their severity of COVID-19, and could not linked their information with secondary cases.

## Definitions

A source case is a person diagnosed with COVID-2019 a close contact person has made close contact with. Close contacts may have made contact with one or more patients.

The diagnosis of SARS-CoV-2 infection was made, according to the 6^th^ National Criteria for Diagnosis of COVID-2019 in China^14^. As the study participants were all close contacts, a COVID-2019 case was referred to a person who had both a positive result for the virus’ nucleic acid and symptoms and/or abnormal radiological/laboratory findings before, during or even after the 14 days of quarantine. Asymptomatic infection must have not clinical symptoms, must be positive for the virus’ nucleic acid, and have or be free of radiological and/or laboratory alterations that indicate viral infection.

Fever was defined as an axillary temperature of 37.5°C or above. Severity of the disease includes 5 categories: asymptomatic, mild, moderate, severe and critical. Mild cases were those who had mild symptoms but no radiological alterations. Moderate cases are those who had both symptoms and radiological alterations. Severe cases were those who had any of the following: breathing rate ≥30/min, or oxygen saturation level ≤93% at rest, or oxygen concentration level PaO^2^/FiO^2^ ≤ 300mmHg (1mmHg=0.133kPa), or lung infiltrates >50% within 24 to 48 hours. Critical cases are those who had respiratory failure requiring mechanical ventilation, septic shock, or multiple organ dysfunction/failure.

The mode of contact was classified into 5 categories: public transport vehicles, healthcare settings, households, multiple, and others. Tourists in the ship cruise were put in a special exposure group called “Dream Cruise”. The multiple contact includes those who were exposed to more than one mode of contact (e.g. household and public transport vehicles).

### Diagnosis of RT-PCR test, radiological and blood examination

Throat swab samples were collected by trained CDC staff and transported and stored in −70 L refrigerators in biological safety level 2 laboratories. Samples of cluster cases were also sent to China CDC for re-examination. RT-PCR testing was performed by qualified staff and results were identified through open reading frame 1ab (ORF1ab) and nucleocapsid protein (N) in accordance with the protocol established by China CDC^13^. Details on laboratory processes are provided in Appendix 3. Radiological and blood examinations were conducted in tertiary hospitals designated for treating COVID-19 patients according to national standards^14^.

### Statistical analysis

The infection rate was estimated by dividing the number of diagnosed cases with the number of close contact persons and compared among different contact groups. Categorical variables were described in number and percentage (%), and continuous variables in median and interquartile range (IQR). Differences in proportions were tested by using the *χ*^2^ test. Univariate and multivariable logistic regressions were performed to adjust for potential factors that may affect the risk of developing COVID-19, and odds ratio (OR) and 95% confidence interval (95% CI) were estimated.

Analyses were all performed with the SAS software (version 9.4 for Windows, SAS Institute, Inc., Cary, NC, USA). Statistical tests were two-sided, and *P* values of less than 0.05 were considered to indicate statistical significance.

### Ethics Approval

Ethics approval was obtained from the Ethics Committee of Southern Medical University. Data collection and analysis of close contacts and source cases were also required by the National Health Commission of the People s Republic of China to be part of a continuing public health outbreak investigation. Written informed consent was waived in light of the urgent need to collect data.

## Results

### Baseline characteristics of close contacts

By the end of the Mar 6, 2020, all the 4950 close contacts were enrolled. Males accounted for 2484 (50.2%). The median (IQR) age was 38.0 years and 783 (15.8%) were under 18 years (Table 1). Exposure in public transports was the commonest type of close contact. On average, 2.4 PCR tests were performed for each person. 129 (2.6%) cases were identified with 8 (6.2%) being asymptomatic throughout and 5 (3.9%) being clinically severe or critical.

**Table 1.**
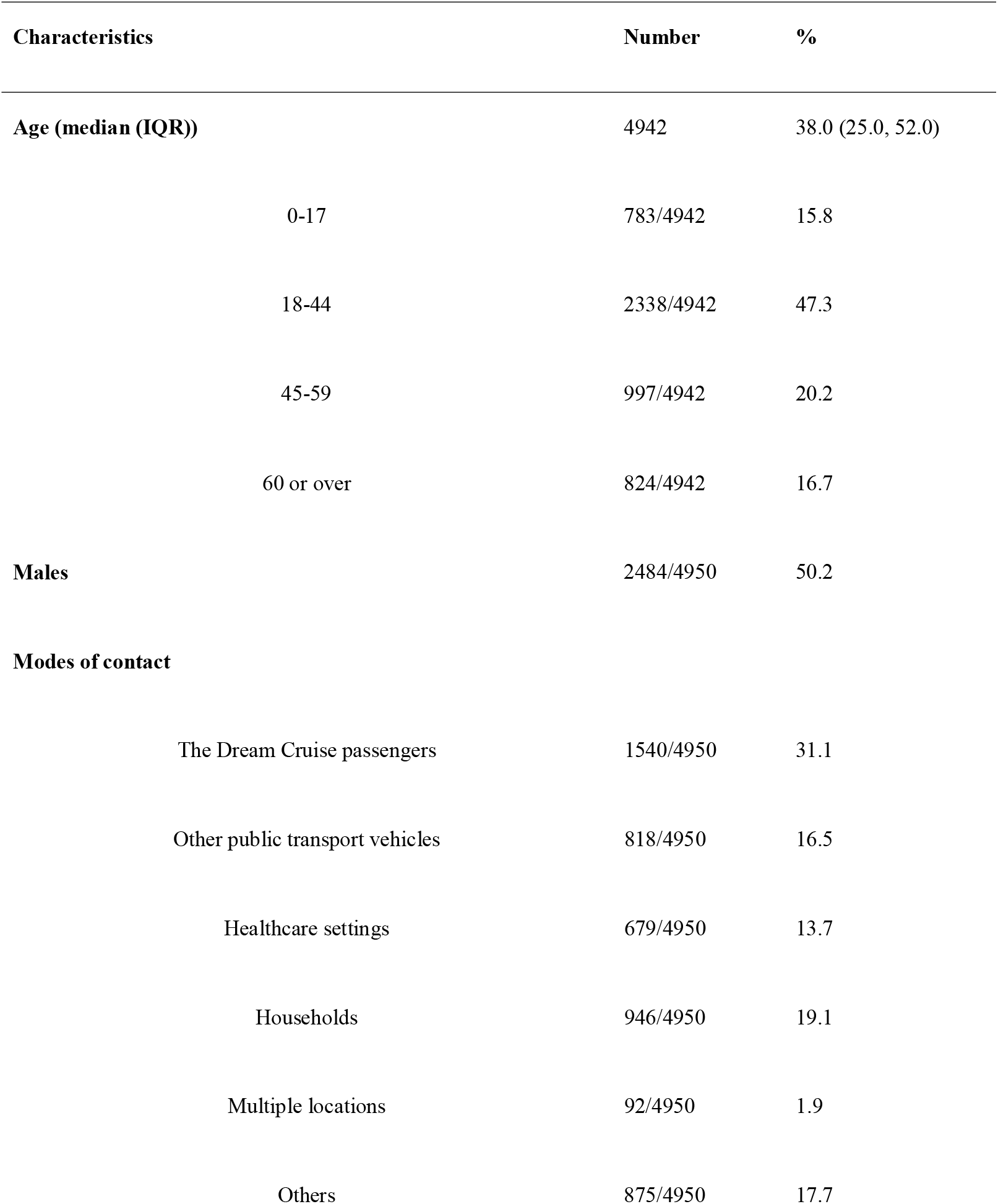
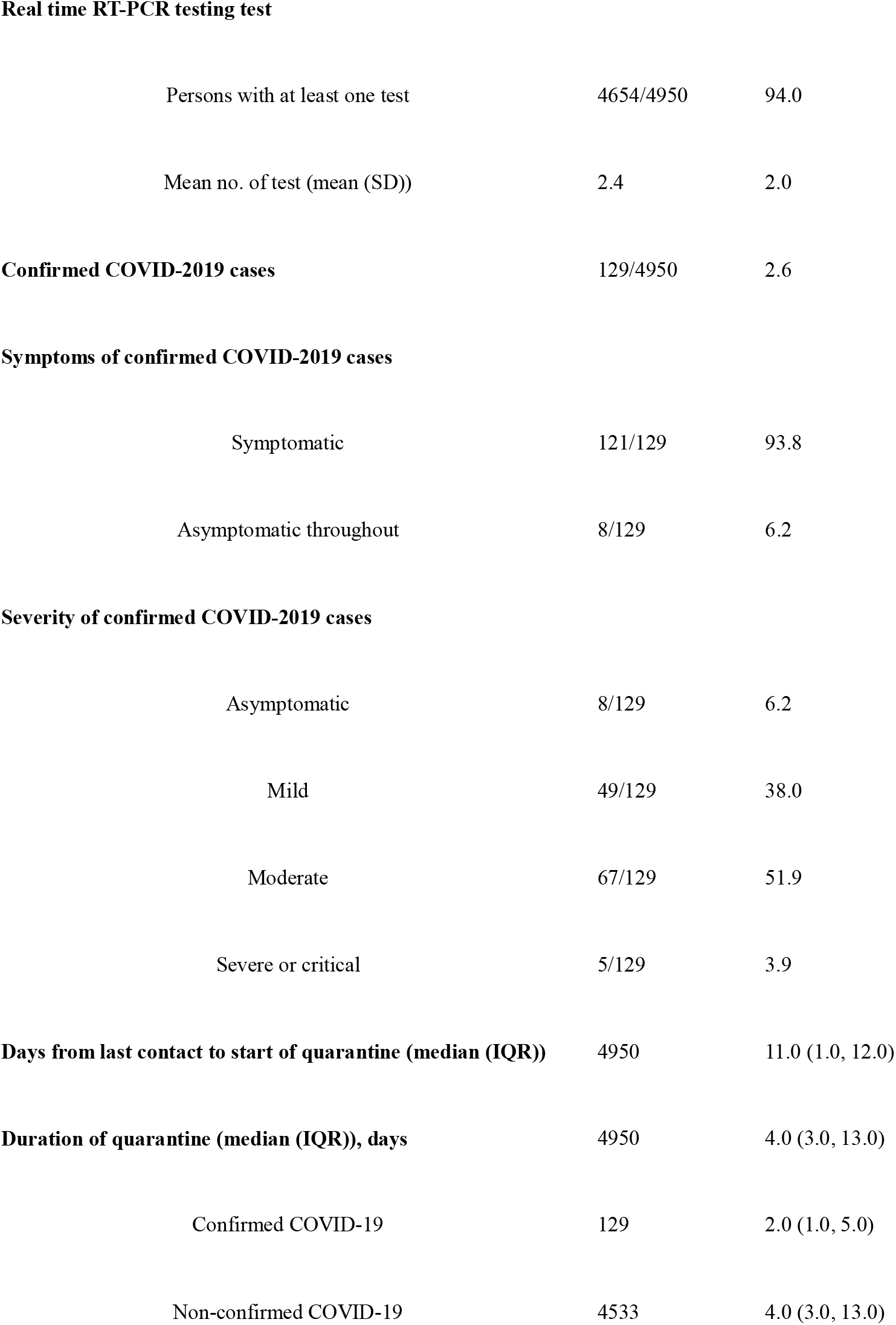
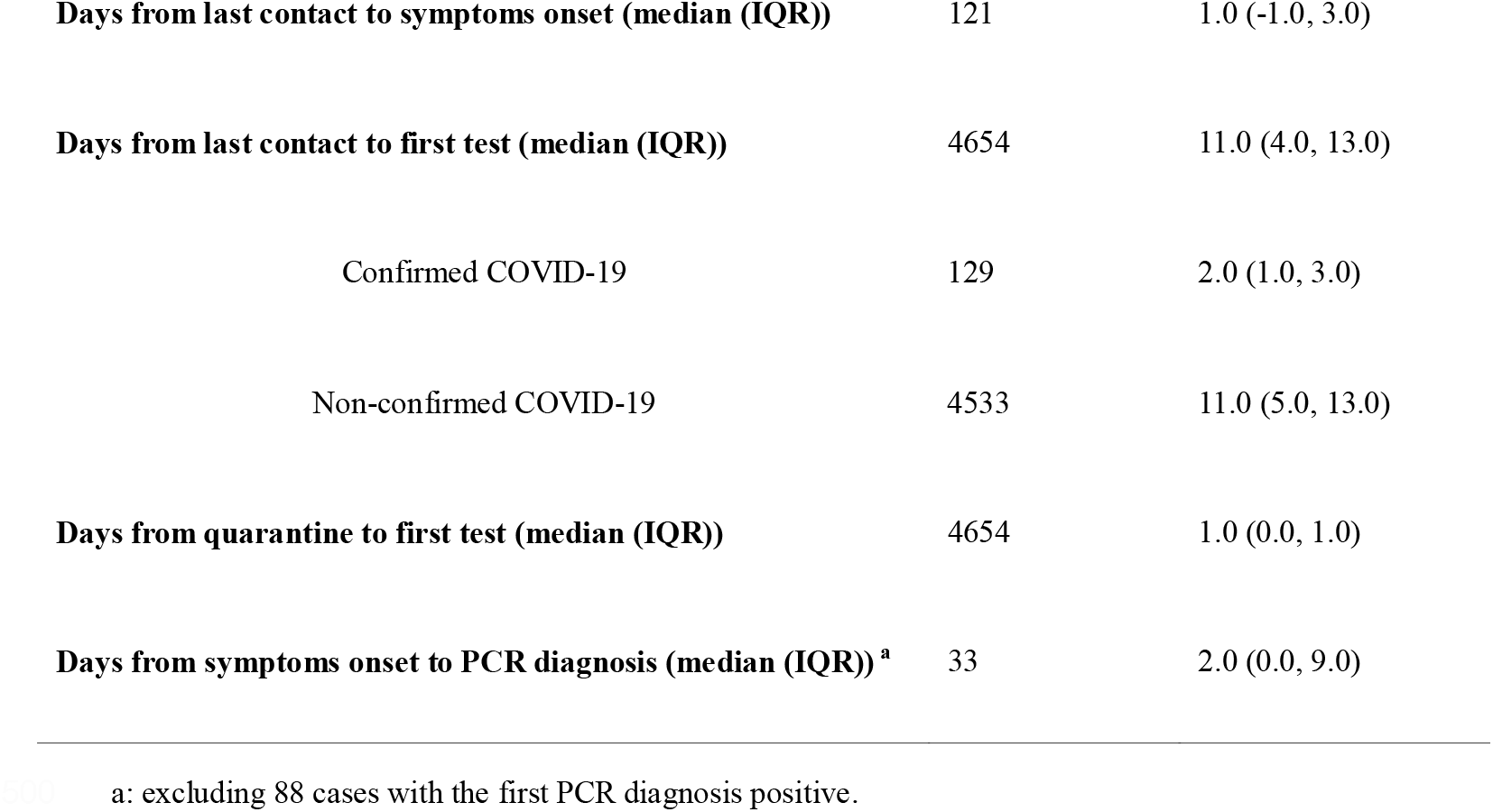
Baseline characteristics of 4 950 Persons with Close Contact with Confirmed COVID-2019 Cases, Guangzhou, China.

| Characteristics | Number | % |
| --- | --- | --- |
| **Age (median (IQR))** | 4942 | 38.0 (25.0, 52.0) |
| 0-17 | 783/4942 | 15.8 |
| 18-44 | 2338/4942 | 47.3 |
| 45-59 | 997/4942 | 20.2 |
| 60 or over | 824/4942 | 16.7 |
| **Males** | 2484/4950 | 50.2 |
| **Modes of contact** |  |  |
| The Dream Cruise passengers | 1540/4950 | 31.1 |
| Other public transport vehicles | 818/4950 | 16.5 |
| Healthcare settings | 679/4950 | 13.7 |
| Households | 946/4950 | 19.1 |
| Multiple locations | 92/4950 | 1.9 |
| Others | 875/4950 | 17.7 |
| **Real time RT-PCR testing test** |  |  |
| Persons with at least one test | 4654/4950 | 94.0 |
| Mean no. of test (mean (SD)) | 2.4 | 2.0 |
| **Confirmed COVID-2019 cases** | 129/4950 | 2.6 |
| **Symptoms of confirmed COVID-2019 cases** |  |  |
| Symptomatic | 121/129 | 93.8 |
| Asymptomatic throughout | 8/129 | 6.2 |
| **Severity of confirmed COVID-2019 cases** |  |  |
| Asymptomatic | 8/129 | 6.2 |
| Mild | 49/129 | 38.0 |
| Moderate | 67/129 | 51.9 |
| Severe or critical | 5/129 | 3.9 |
| **Days from last contact to start of quarantine (median (IQR))** | 4950 | 11.0 (1.0, 12.0) |
| **Duration of quarantine (median (IQR)), days** | 4950 | 4.0 (3.0, 13.0) |
| Confirmed COVID-19 | 129 | 2.0 (1.0, 5.0) |
| Non-confirmed COVID-19 | 4533 | 4.0 (3.0, 13.0) |
| **Days from last contact to symptoms onset (median (IQR))** | 121 | 1.0 (-1.0, 3.0) |
| **Days from last contact to first test (median (IQR))** | 4654 | 11.0 (4.0, 13.0) |
| Confirmed COVID-19 | 129 | 2.0 (1.0, 3.0) |
| Non-confirmed COVID-19 | 4533 | 11.0 (5.0, 13.0) |
| **Days from quarantine to first test (median (IQR))** | 4654 | 1.0 (0.0, 1.0) |
| **Days from symptoms onset to PCR diagnosis (median (IQR))<sup>a</sup>** | 33 | 2.0 (0.0, 9.0) |

a: excluding 88 cases with the first PCR diagnosis positive.

The 4950 close contacts were quarantined for an average of 4.0 days, with 2.0 days for cases and 4.0 days for non-cases (Table 1 and Figure 1). In 20 persons, quarantine was unnecessary as they last contacted a patient 14 days ago and were free of symptoms and PCR test negative at the time they were identified. In 340 persons, quarantine was longer than 14.0 days because the PCR tests were delayed (Figure 1). There was on average 1.0 day from the start of quarantine to the first PCR testing, suggesting a slight delay in collecting samples for laboratory diagnosis (Table 1). PCR diagnosis was made within 14 days of quarantine for all 129 cases but two for whom it was on the 16^th^ day; all the 8 asymptomatic cases were diagnosed within 10 days of quarantine (Figure 1).

**Fig 1.**
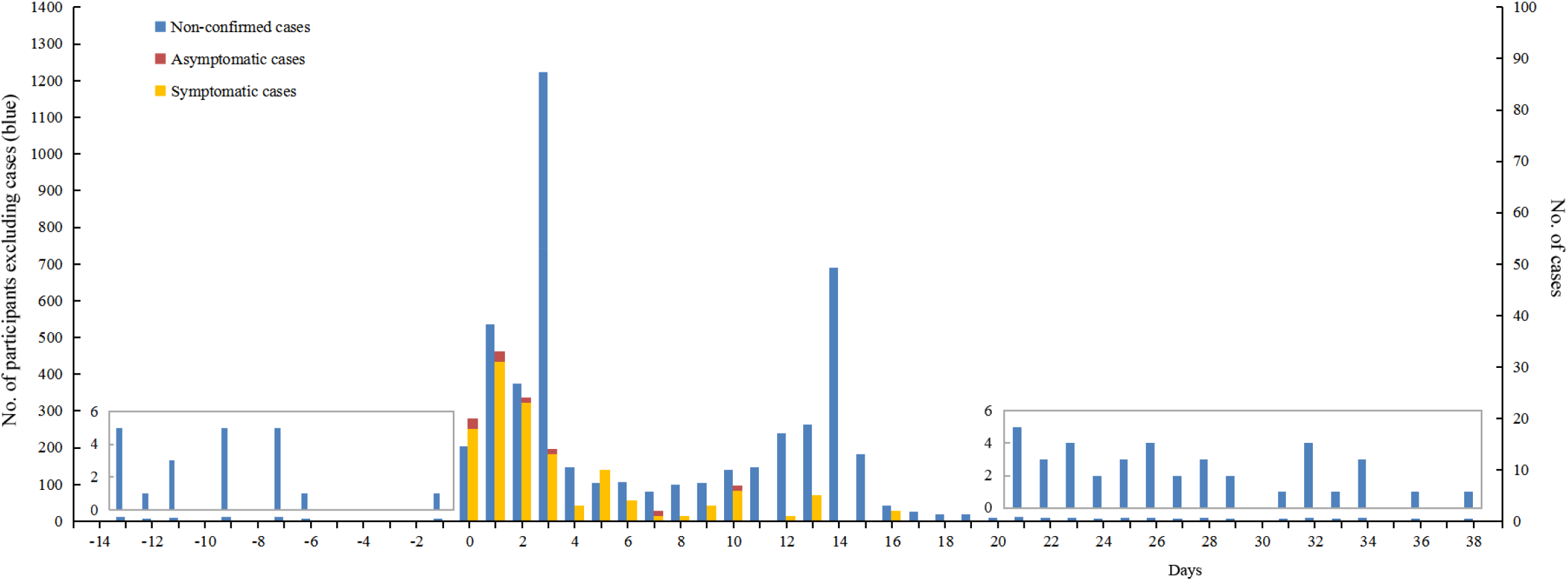
Distribution of 4950 close contact persons by the number of days from start of quarantine to PCR diagnosis or release from quarantine and infection status (day 0 is the day quarantine starts)

There were on average 11.0 days from the last contact to the start of quarantine (Table 1), suggesting quarantine could in theory start 11.0 days earlier than it actually did. The delay from the last contact to quarantine was on average 1.0 day with over 3 days for 11 cases (Table 1 and Figure 2). In symptomatic cases, there was on average 1.0 day from the last contact to symptoms onset, with 31 cases having already developed symptoms before the last contact and 22 cases over 3 days after the last contact (Table 1 and Figure 2). In 33 cases for whom the date of symptoms onset was clear and the first PCR test was negative, we estimated that there were on average a delay of 2.0 days from symptoms to first PCR positivity and in 22 (66.7%) cases symptoms appeared 7.0 days prior to PCR positivity (Figure 2).

**Fig 2.**
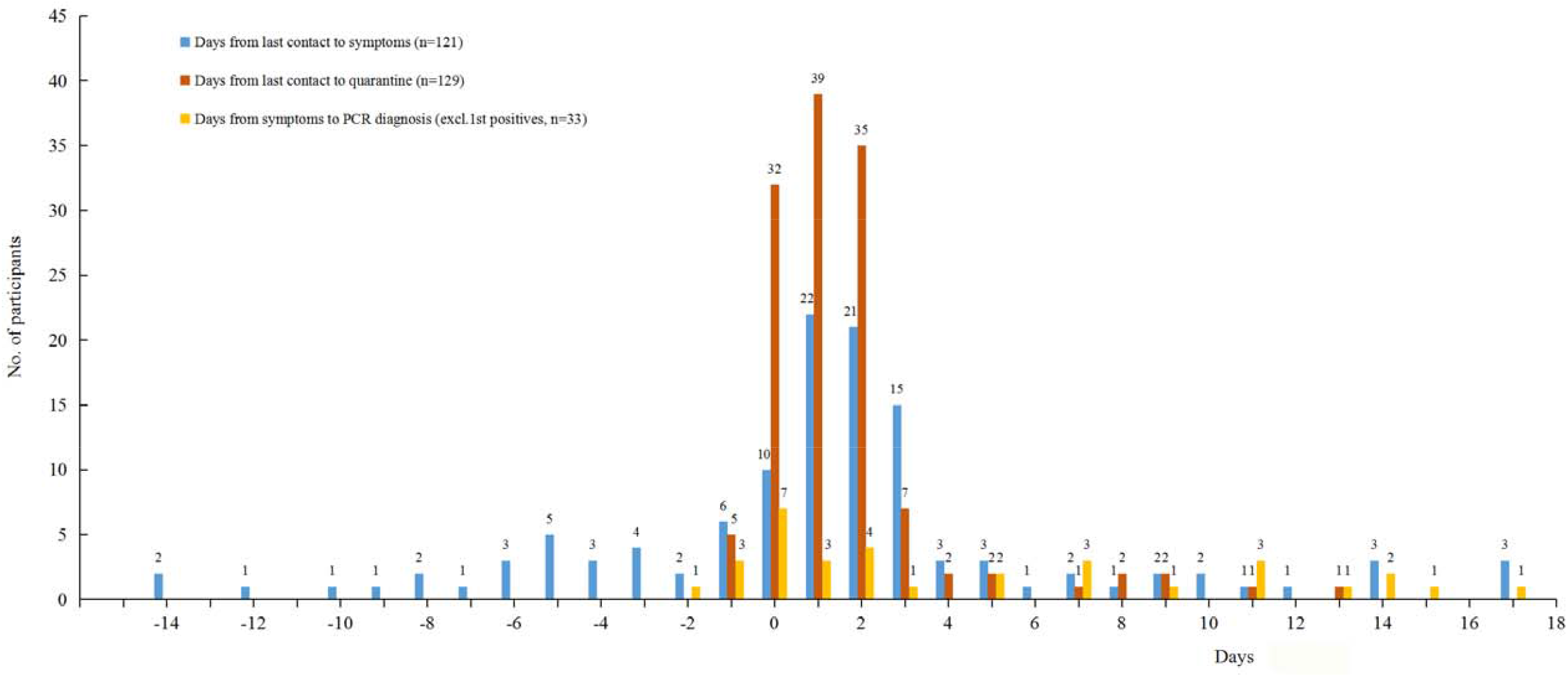
Distribution of days from last contact to symptoms onset and to start of quarantine and days from symptoms onset to PCR diagnosis.

### Mode of contact and risk of transmission

The age of close contacts was linearly associated with an increasing risk of getting infected after close contact with source patients (Table 2). The incidence was 1.8%, 2.2%, 2.9%, and 4.2% respectively for 0-17, 18-44, 45-59, and 60 or above age-groups (*P*=0.0016 for trend). Females seemed as likely as males to catch the infection after close contacts with patients (*P*=0.1202).

**Table 2.**
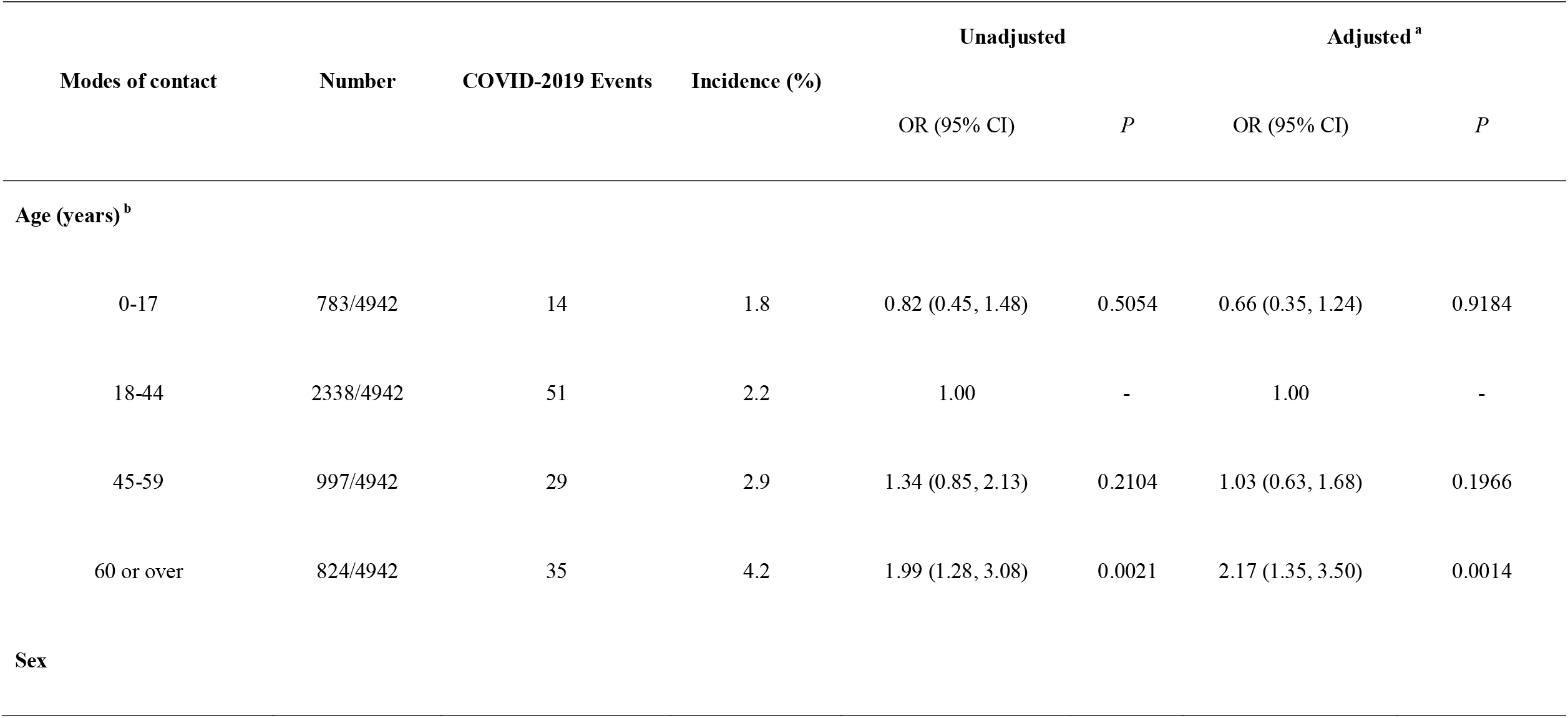
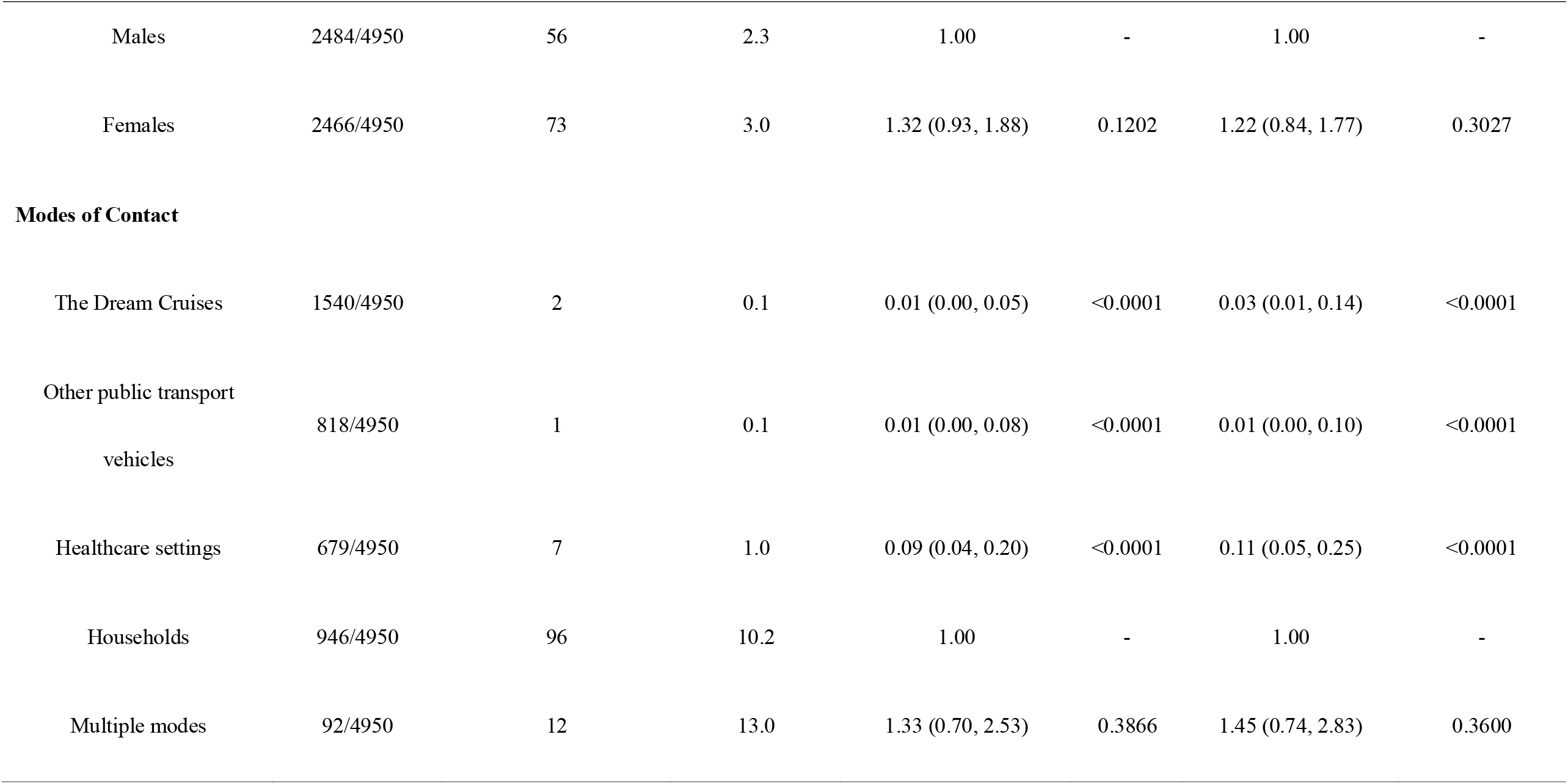
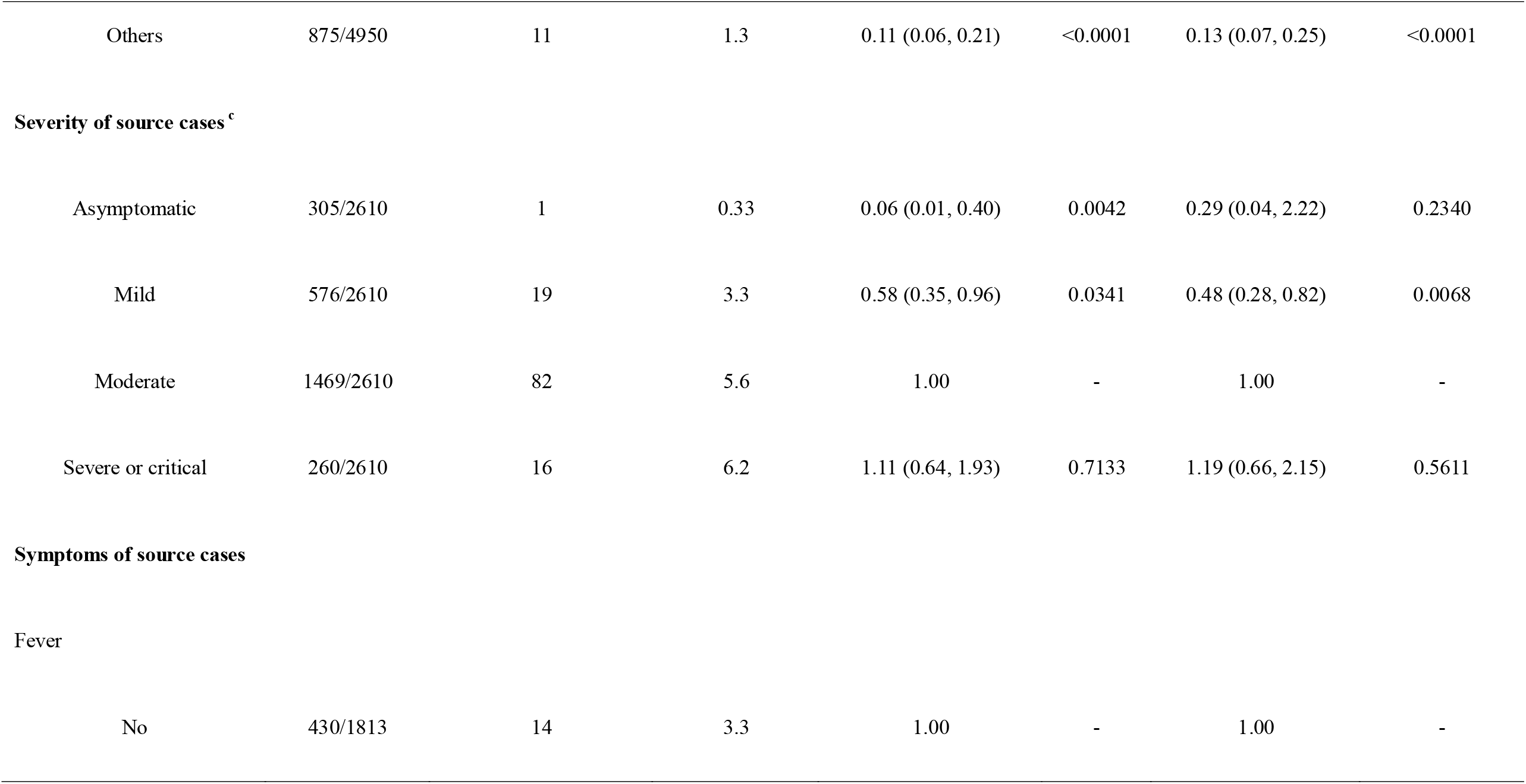
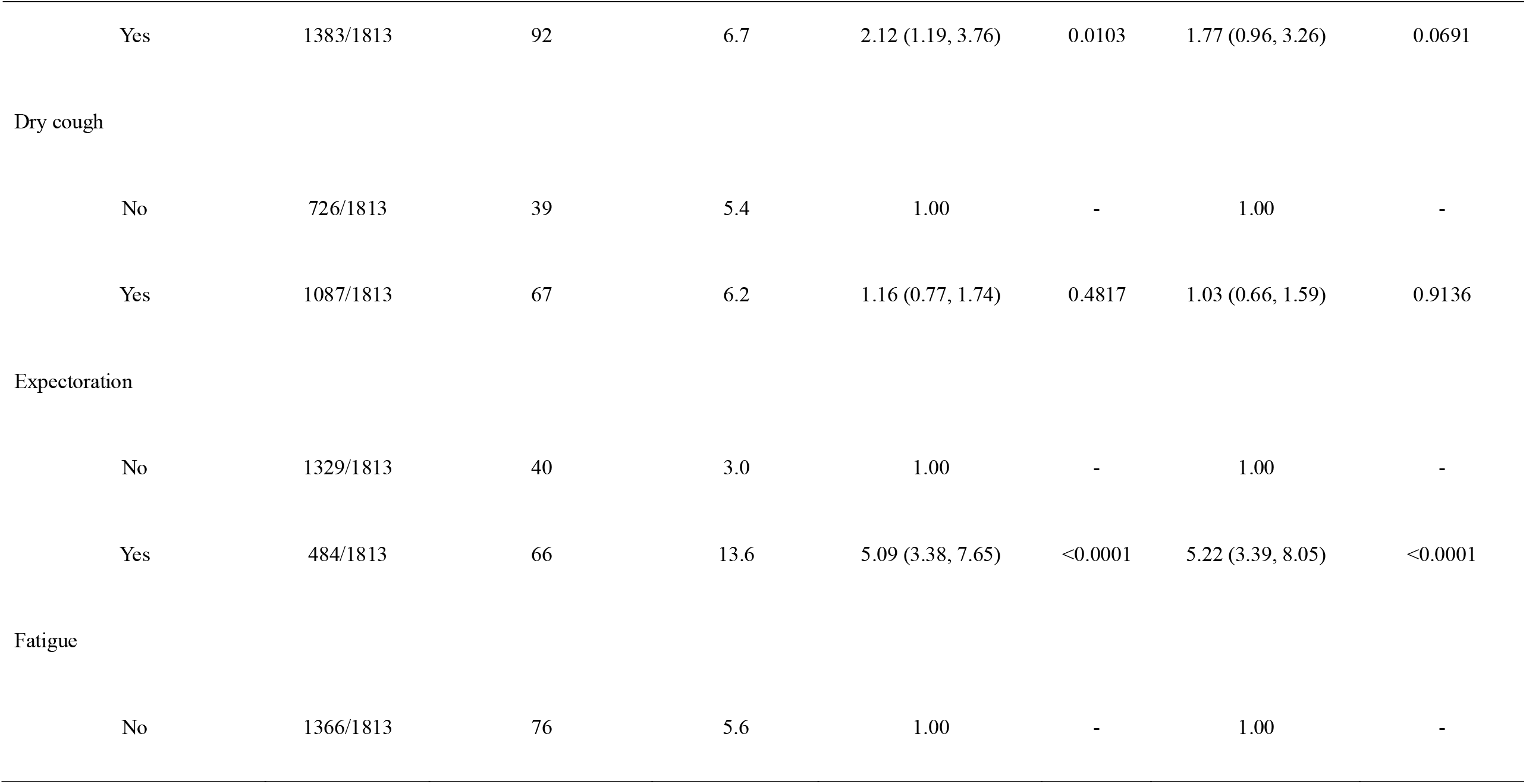
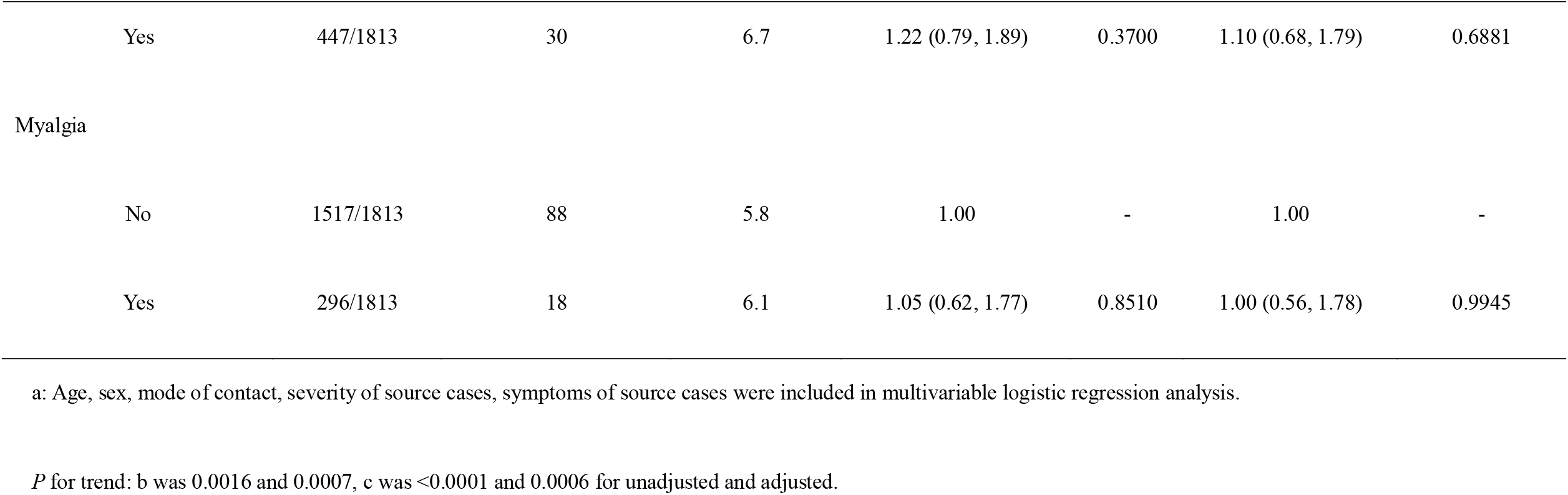
Modes of contact and risk of transmission among 4 950 Close Contact Persons.

Among different modes of contact, household contacts and multiple contacts (with 70% including household contacts) were most dangerous in catching the infection and associated with an incidence of COVID-19 10.2% and 13.0%, respectively (Table 2). Healthcare settings contacts and public transport vehicles, the other two common forms of contacts, were associated with a risk of 1.0% and 0.1%, which were only about 10% and 1%, respectively, of the risk of household contacts (*P*<0.0001).

Furthermore, clinically more severe patients were more likely to pass the infection to their close contacts than less severe ones (*P*<0.0001 for trend). Asymptomatic infection is least likely to pass on the infection, with a chance of 33 per 100,000 contacts. Mild and moderate infections could increase the risk to 3.3% to 5.6%, and severe and critical infections to 6.2%. Manifestation of some symptoms in source patients was also associated with an increased risk of infection in their close contacts. For example, fever could increase the risk by over 100% (*P*=0.0103) and expectoration by 400% (*P*<0.0001), whereas cough, fatigue and myalgia did not statistically significantly increase the risk (*P*>0.3700). In addition, a higher frequency of contact and greater number of patients contacted were highly associated with household contacts and thus were not separately assessed (Table 1S).

The above conclusions remained unchanged and statistically significant in multiple regression analyses which included age, sex, mode of contact, severity of source patients and expectoration included in the models (Table 2).

### Comparison of source cases and secondary cases

We compared the characteristics between secondary cases and source cases they contacted with to see whether they may differ in the severity of the infection. Among 129 secondary cases, source cases were identifiable only for 121 cases. As compared with their 69 source cases, the 121 secondary cases were in general clinically milder and were less likely to have such common symptoms as fever, cough, expectoration, fatigue, myalgia and diarrhea (*P*<0.05). Secondary cases are also less likely than source cases to demonstrate radiological and laboratory alterations related to the infection (*P*<0.001). Most of the differences between them were both clinically important and statistically significant.

The clinical differences between source and secondary patients might be due to the fact that secondary cases were diagnosed earlier and the disease is milder at the early stage than source cases. To exclude this possibility, we also compared source cases with secondary cases who were diagnosed before the time of quarantine and not supposed not be early-stage patients. The conclusion remained unchanged (Table 2S).

### Validity of PCR for Diagnosis

Among 4950 close contacts, 4653 completed at least one RT-PCR testing. If a person has no symptoms and the PCR test was negative, further testing continued to be arranged within 48 hours till he was diagnosed with the infection or released free of the infection from quarantine. The series of testing in the same persons allowed us to estimate the sensitivity and specificity of the PCT testing. The results were shown in (Table 3S). In brief, the sensitivity was only 71.9% for the first testing and increased to 92.2% by the second testing, to 96.9% by the third testing, and to 100.0% by the sixth testing. In contrast, the first testing achieved a specificity 99.96%, which was reduced by less than 0.1% by further testing.

**Table 3.**
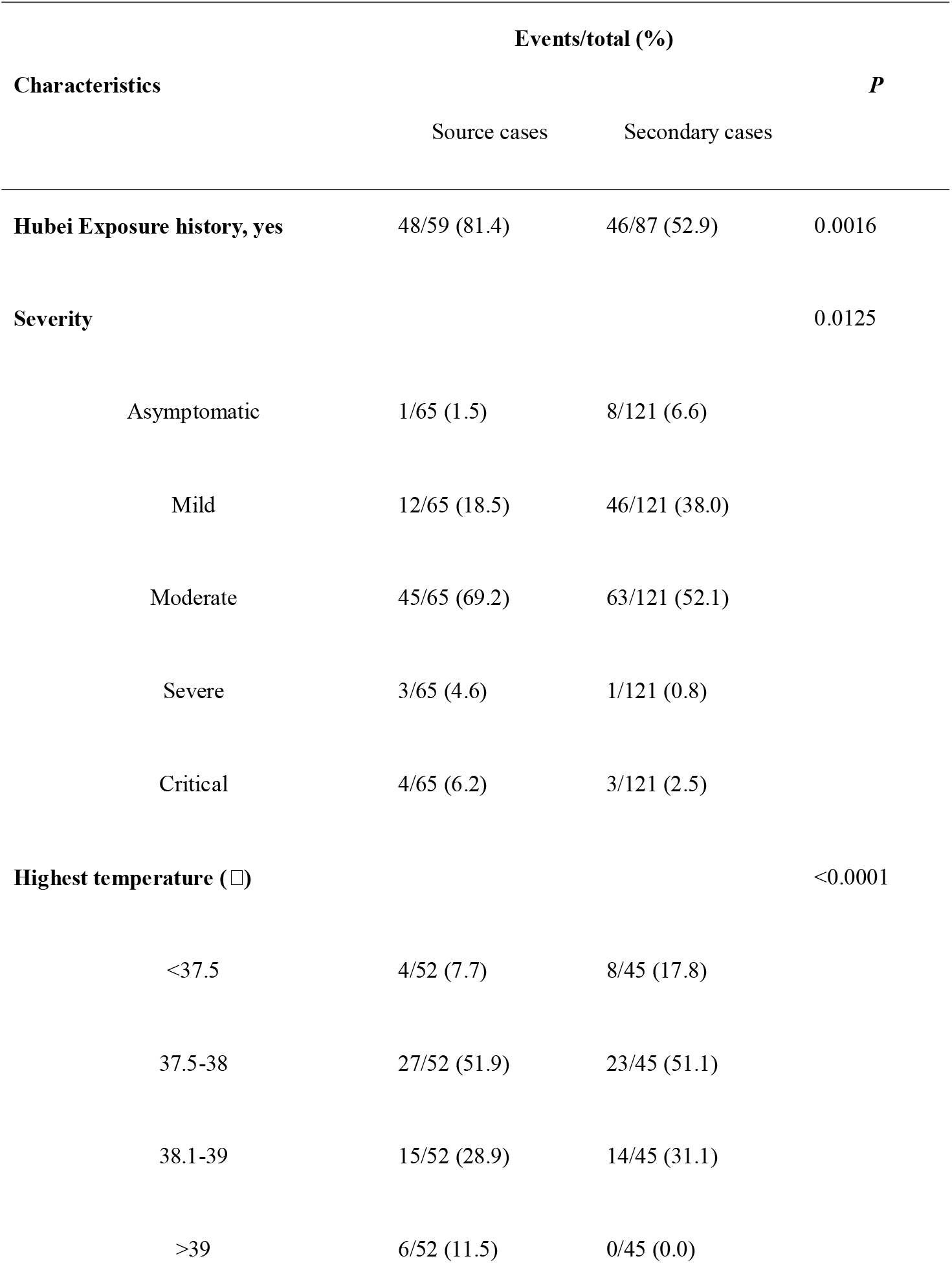
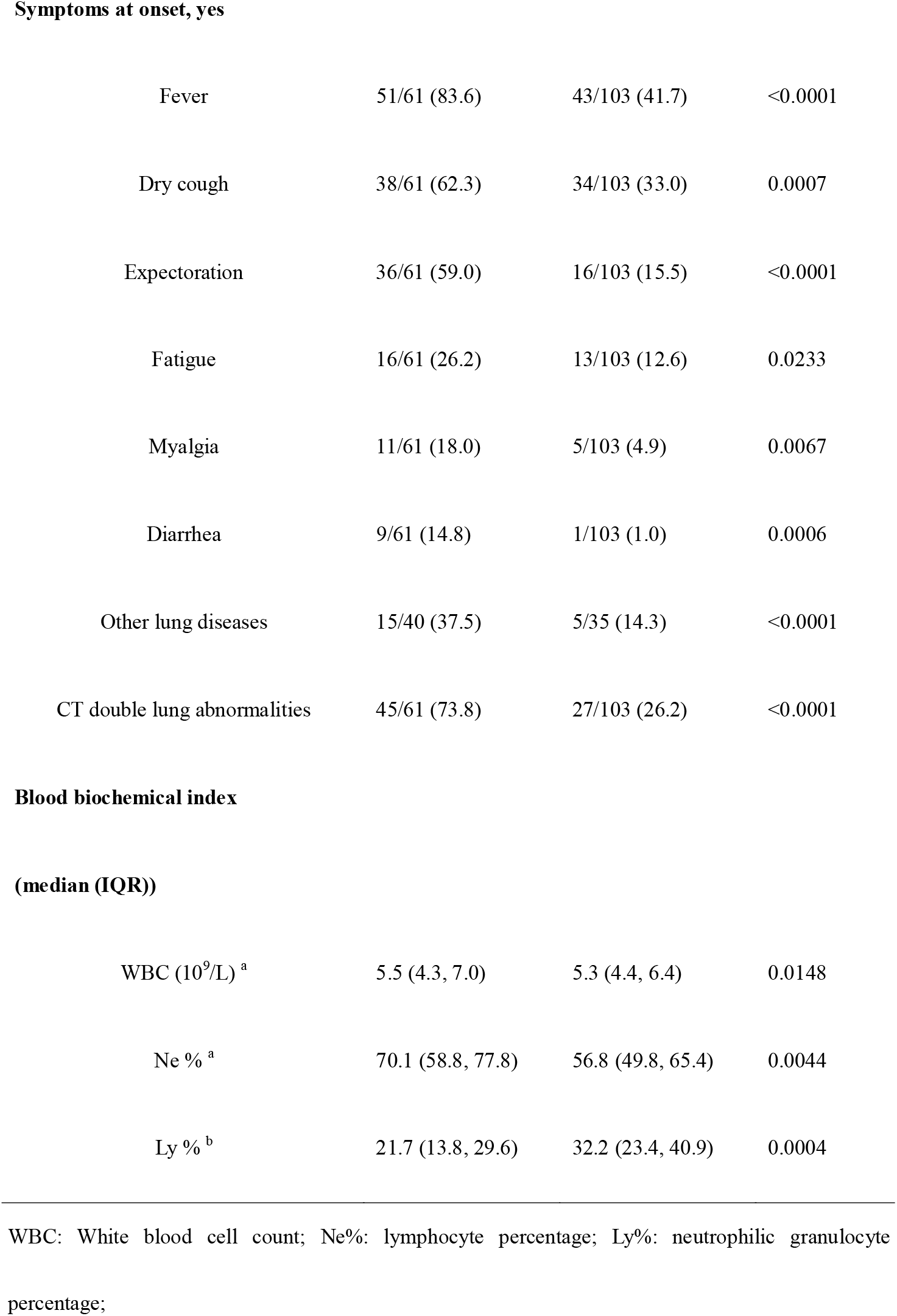
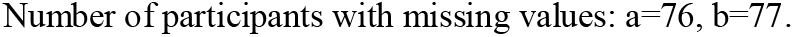
Comparison of clinical, radiological and laboratory characteristics of COVID-2019 infection between 69 source cases and 121 secondary cases.

## Discussion

Between January 13, and March 6, 2020, 4950 close contacts of confirmed cases were enrolled in Guangzhou, which is a city with large confirmed cases of COVID-19 outside Hubei province in China. Here we evaluated the modes of contact and risk of transmission among close contacts, provides insights into transmission and control of COVID-19. To our knowledge, this study is the largest prospective cohort data of close contacts with COVID-19.

Our study provided further evidence that the older aged contacts and household contacts were more likely to be infected^3,4,7,15^. The incidence of asymptomatic and mild infections was high (57/129), and the risk of transmission increased as the symptoms of source cases worsen, with range from 0.33% (asymptomatic) to 6.2% (severe and critical). The symptomatic cases with expectoration symptom had a higher transmission capacity. The results provide the evidentiary foundation for evaluating control measures, and guiding the global response.

Household contacts characteristics have been discussed in previous studies^3,7^, which were the source of person-to-person transmission evidence. And our study further confirmed that due to contact frequently of household, it was considered as a high-risk factor for COVID-19 transmission. The incidence of household contacts estimated 10.2% in our study, and in other study out of Hubei Province was 14.9 %^16^, which is consistent with current understanding of COVID-19 transmission. However, other modes of contact have been less reported for guiding persons self-protection and government for strengthened control measures. In our study, 1540 close contacts were Dream Cruise passengers, and the infected incidence was 0.1% (2/1540), which was estimated low. This result was consistent with previous research, and Nishiura H^17^ estimated the incidence of infection with COVID-19 on a cruise ship, called Diamond Princess, and the risk of infection among passengers contact occasionally was considered to be very limited.

However, the food service workers had a high infected incidence in the cruise ship^18^ largely due to their frequent contact to others and had high chance to inhale droplet spread.

The risk transmission of other public transport vehicles and healthcare settings was estimated low, which were only about 1% and 10% of the risk of household contacts (*P*<0.0001). It suggests that control personnel density on public places and distance oneself from others are very effective prevention and control measures^19^. Giving that public transport vehicles was at low risk to infected COVID-19, and it is considered feasible to take public transportation when returning to work or school on the premise of low personnel density.

The proportion of close contacts confirmed with asymptomatic and mild were high with 44.2 % (57/129). Chowell^20^ estimated asymptomatic proportion was at 17.9% among 700 infected individuals on Diamond Princess, and Miyama T^21^ estimated at 30.8% among 13 Japanese evacuees from Wuhan City. Taking the results from several studies into account, Chowell^20^ thinks that asymptomatic or mild cases combined represent about 40% to 50% of all infections. It was consistent with the results of this study.

Given the large proportion of asymptomatic and mild infections, we are concerned about the rate at which they infect others. Wendtner^22^ showed that patients with COVID-19 had high levels of the virus in throat swabs early in their illness, when their symptoms were mild. But no study had reported the infection risk of asymptomatic and mild cases to others, and asymptomatic infections might be seeding new outbreaks^23^. In our study showed that as severity increases, the risk of transmission increases in COVID-19 patients. Only 1 (1/305, 0.33%) and 19 (19/576, 3.3%) close contact was infected by asymptomatic and mild source case, and it suggested the limited transmission capacity in asymptomatic and mild cases. The symptomatic cases with expectoration symptom had a higher transmission capacity. These might be associated with more viral load of SARS-CoV-2 in patients with severe symptoms^24^.

Given the current evidence, due to asymptomatic cases have limited transmission capacity, then the primary surveillance and control measures should focus on symptomatic contacts. On the other hand, though a person with asymptomatic or mild symptoms may not easy to spread SARS-COV-2, and had a low probability to infect other. While, we should also be alert for incubation transmission^25^. Asymptomatic and mild patients might not aware of their infection and therefore not isolate themselves or seek treatment, or they might be overlooked by health-care professionals and thus unknowingly transmit the virus to others. Due to the imperfect sensitivity of the PCR test (Table 3S), some asymptomatic contacts may be missed^26,27^. Thus, based our evidence, two times or more PCR tests were recommended to ensure that almost all patients could to be diagnosed.

Previous studies suggested that compared with patients initially infected with SARS-Cov-2 in Wuhan City, the symptoms of patients in out of Wuhan are relatively mild^28,29^. And a research reported that the symptoms of imported cases (n=15) were severe than those of secondary cases (n=17)^30^, but due to the small sample size, it may be necessary to verify the phenomenon. Thus, our study compared the severity of symptom between sources cases and their secondary cases. And the severity of clinical symptoms onset was more severe to source cases compared to secondary cases (*P*<0.001). It may be related to the higher Hubei exposure history of source cases (20/33 vs. 21/37) than secondary cases. This phenomenon was also apparent during the transmission of MERS-CoV^31^.

Our study has some notable limitations. Firstly, we have not the data to show the prognosis of disease. Because many patients remained in the hospital and the outcomes were unknown at the time of data cutoff, we censored the data regarding their clinical outcomes and thus entire course of the disease cannot be fully demonstrated. Secondly, we used logistic regression analysis instead of cox proportional hazards model, because of the low incidence (2.6%) of COVID-19 among close contacts. In addition, by the end of the cohort, there were 245 close contacts remaining quarantines, but they were not likely to become COVID-19 cases, thus there was no censored data. Thirdly, there may be a recall bias of the symptoms at onset among source cases and secondary cases.

In conclusion, our cohort study showed that the proportion of asymptomatic and mild infections account for almost half of the confirmed cases among close contacts. The household contacts were the main transmission mode, and clinically more severe cases were more likely to pass the infection to their close contacts. In general, the secondary cases were clinically milder than those of source cases. The results provide the evidentiary foundation for evaluating control measures, and guiding the global response.

## Data Availability

The raw/processed data required to reproduce these findings cannot be shared at this time as the data also forms part of an ongoing study.

## Supplementary Material

Table 1S. Modes of contact and risk of transmission among 4 950 Close Contact Persons;

Table 2S. Comparison of clinical, radiological and laboratory characteristics of COVID-2019 infection between 36 source cases and 49 secondary cases;

Table 3S. Sensitivity, specificity, and positive and negative predictive values of sequential nucleic acid tests of throat swabs (n=4653);

Figure 1S. Distribution of 1540 Dream Cruises close contact persons by the number of days from start of quarantine to PCR diagnosis or release from quarantine and infection status (day 0 is the day when quarantine starts);

Appendix 1: The full detail of close contacts;

Appendix 2: COVID-19 Confirmed Case Investigation Form;

Appendix 3: Details regarding laboratory confirmation processes.

